# Proof of Concept: detection of cell free RNA from EDTA plasma in patients with lung cancer and non-cancer patients

**DOI:** 10.1101/2022.08.12.22278721

**Authors:** Kristin E. Mullins, Chamindi Seneviratne, Amol C. Shetty, Feng Jiang, Robert Christenson, Sanford Stass

## Abstract

Nucleic acid sequencing technologies have advanced significantly in recent years, allowing the development of liquid biopsies as new means to detect cancer biomarkers and cancer heterogenicity. Most of the assays available clinically focus on cell free DNA, however, cell free RNA (cfRNA) is also present and has the potential to complement and improve cancer detection especially in cancers like lung cancer, which are usually only diagnosed at late stages and therefore have poor long-term survival outcomes. Remnant EDTA plasma was collected from lung cancer patients and non-cancer individuals at the University of Maryland Medical Center. RNA was extracted and processed for next generation sequencing with a tagmentation-based library preparation approach. cfRNA was successfully extracted and sequenced from 52 EDTA-treated plasma samples with volumes as low as 1.5 mL. This quantity was sufficient to prepare libraries with the length of libraries averaging from 264bp to 381bp and resulted in over 2.2 to 3.6 million total sequence reads respectively. Sequential dilution of cfRNA samples from healthy individuals indicated that the starting cfRNA concentration influenced the detection of differentially expressed genes. This proof-of-concept study provides a framework for screening cfRNA for identifying biomarkers for early detection of lung cancer (and other cancers), using minimal amounts of samples (1.5 ml) from standard EDTA 3-mL collection tubes routinely used for patient care. Further studies in large populations are required to establish limit of detection and other parameters including precision, accuracy, sensitivity, and specificity, to standardize this method.

## INTRODUCTION

Lung cancers make up the second most common cancer diagnosis in the United States and leading cancer diagnosis world-wide. Further, lung cancers are the leading cause of cancer-related deaths due to the late-stage diagnoses with 5-year survival rates of 10-15%(1, 2). While advancements in detection and treatment have led to earlier detection and longer survival, the majority of patients are not diagnosed until symptomatic and current diagnoses rely heavily on chest radiographs (1,3). Furthermore, current biomarkers, such as carcinoembryonic antigen (CEA), are not indicated for screening but rather as indicators of prognosis and disease reoccurrence(4). For instance, Cancer-SEEK for combined analysis of circulating DNA mutations and proteins can diagnose eight common cancer types, including lung cancer. However, it has only 40% sensitivity for early-stage lung cancer. Therefore, biomarkers for early and non-invasive cancer detection are lacking for lung cancer and other late-stage diagnosed cancers.

Recent advances in nucleic acid sequencing technology have had large implications for the clinical lab and for early detection and monitoring of cancers. Many of the recent developments have involved peripheral blood liquid biopsies, which has the advantage of being non-invasive and avoids complications associated with traditional biopsies(5-7). Additionally, unlike molecular techniques using traditional biopsies, liquid biopsies have the advantage of real-time detection, and full tumor profiling of cancers with significant heterogenicity depending on the region of the tumor (7, 8) allowing for clinicians to make better informed decisions regarding treatment protocols and clinical trials.

While liquid biopsies have major advantages compared to traditional techniques, those currently available to clinicians primarily use tumor derived cell free DNA (cfDNA) which poses a number of challenges. Of most importance, cfDNA cannot specify tissue of origin and it is shed into the blood at low levels for slow-growing or small tumors, thus leading to limitations in sensitive, early cancer detection. Because of this major limitation, biomarkers that can complement cfDNA and potentially enhance early detection are sorely needed. (9-11)

Recent reports indicate that cell free RNA (cfRNA) could complement cfDNA in determining the molecular profile of tumors and in advancing early detection. Studies show that cfRNA is released into the circulation by cancer cells; cfRNA could enhance detection of tumors with low burden due to higher expression compared to cfDNA in cancer cells and at higher levels than cfRNA expressed by non-cancerous cells (6, 9, 12).

In this proof on concept study, we focus on the preanalytical testing phase to develop and validate a technique to preserve, extract, and detect cfRNA from small volumes (1.5 mL) of EDTA plasma that can be utilized to develop biomarkers for early-stage lung cancer detection and profiling.

## METHODS

### Sample Collection

We analyzed two sets of samples in this study: Set_1, to test the ability to detect cfRNA, and the second set (Set_2) to explore how the amounts of starting material influence cfRNA detection. For Set_1, whole blood was collected into 3 mL tubes containing EDTA from 20 lung cancer patients (10AC and SSC each) and 32 non-cancer patients (20 smokers without cancer and 12 non-smokers) at the University of Maryland Medical Center. Whole blood samples were processed as described previously, (13) for extraction of remnant plasma and frozen at −80°C until use. Set_2 included plasma and white blood cells (WBC) extracted from the same 8-10ml of whole blood sample containing acid citrate dextrose (ACD) solution, from four healthy volunteers who have provided informed consent to genomic analyses (UMB IRB protocol ID: HP-00060091). Set_2 samples were lysed with red blood cell lysis solution (QIAGEN) to isolate WBC, washed with phosphate-buffered saline for further purification, and preserved in *RNAlater* (ThermoFisher Scientific). Both WBC and plasma samples were stored at - 80°C until use.

### cfRNA processing and sequencing

cfRNA was extracted from 1.5-2 mL of archived plasma for the Set_1 and processed in 200 ul aliquots using the miRNeasy® Serum/Plasma Advanced Kit (Qiagen). The manufacturer’s instructions were followed including the optional DNase treatment steps. cfRNA was eluted twice (10ul followed by 15 ul) from the spin column using 37°C RNase-free water, and frozen at - 80°C until use.

The cellular RNA (cRNA) from WBC was extracted using Macherey-Nagel’s NucleoSpin® miRNA kit and RNA/DNA Buffer kit (Takara Bio USA, Inc. Doral, Fl, USA) according to manufacturer’s recommendations. In Set_2, 4ml of plasma was used for extraction of cfRNA as described above. Next, the total cfRNA (40ul) from each individual was serially diluted with RNase-free water by two-folds to prepare six concentrations of cfRNA per sample.

All total RNA samples (cfRNA and cRNA) were tested for RNA integrity using an Agilent bioanalyzer system and processed for next generation RNA-sequencing with a tagmentation-based library preparation approach consisting of a two-step probe-assisted exome enrichment for cfRNA detection, developed by Illumina. The sequencing workflow is presented in Figure 1. The sequencing alignments were generated by HISAT2. *Samtools* was used to generate alignment statistics.

**Figure 1:**
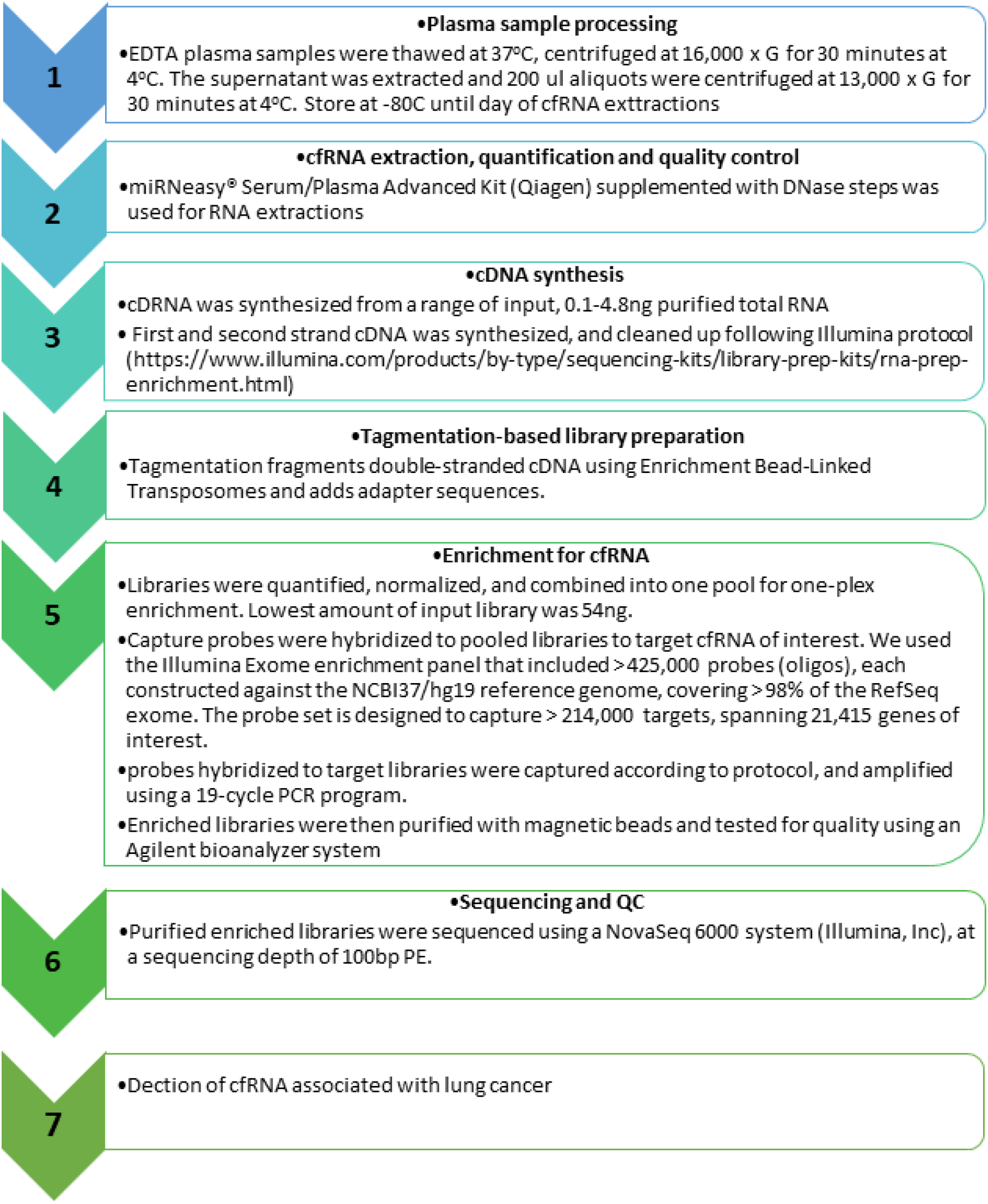
The cfRNA sequencing workflow

## RESULTS

cfRNA was extracted from all 52 plasma samples in Set_1; concentrations ranged from 4-400pg/ul with 1 to 5.3 RNA Integrity Numbers (RIN). These small quantities were sufficient to prepare libraries from each sample as detailed in Figure 1. The length of libraries ranged from 264bp to 381bp and resulted in 223,741,524 to 364,249,966, total sequence reads, respectively. The sequence read distribution in each group are represented in Figure 2. The proportions of exonic, intronic, and intergenic alignments for each group are represented in Supplementary Figure 1.

**Figure 2:**
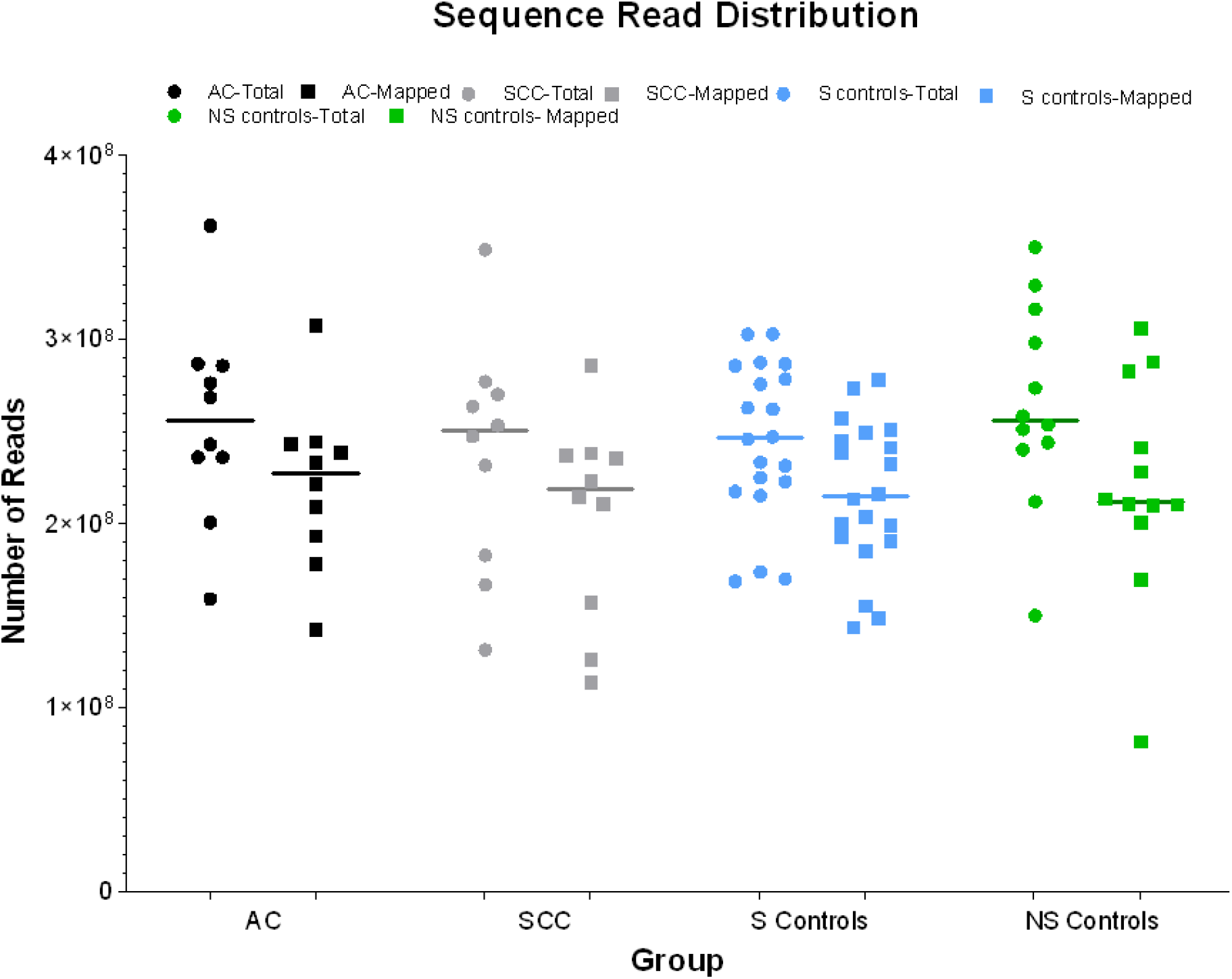
Sequence Read Distribution The total number of reads and the number of mapped reads for all samples in each group of first set of plasma samples. The median line indicates the distribution of total reads or mapped reads within each group. AC-adenocarcinoma; SCC-Squamous cell carcinoma; S controls – smoker controls; NS controls – non-smoker controls.

The average cfRNA concentrations of the six serially diluted plasma samples ranged from 210 to 39pg/ul. The Supplementary Figure 1 presents variability in detecting cfRNA in serially diluted Set_2 samples and cRNA (RIN >7) analyzed by Pearson’s correlation method in R Statistical software. The serially diluted cfRNA samples had 122 to 255 differentially expressed genes (DEGs) compared to the undiluted cfRNA samples from lowest to highest dilutions. The cRNA samples diluted ten folds had an average of 24 DEGs compared to the undiluted cRNA samples. An average of 8,252 (7,173 upregulated and 1,079 downregulated) genes were differentially expressed between undiluted cRNA and cfRNA samples.

**Supplementary Figure 1:**
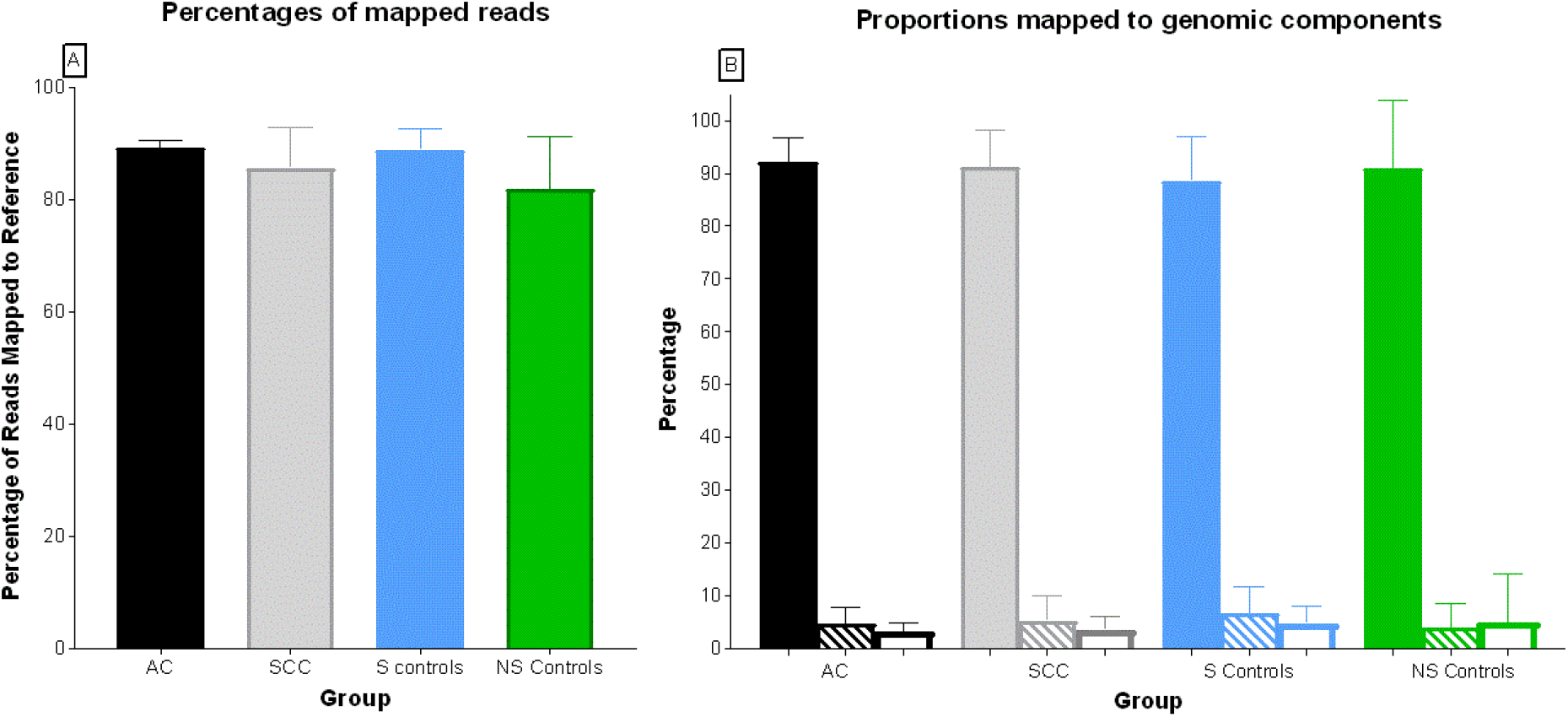
Sequence mapping statistics. (A) Percentages of mapped reads. **(B)** The proportions of reads mapped to different genomic components. Solid bars represent percentage mapped to exonic regions, bars with lines represent reads mapped to intronic regions, and open bars represent reads mapped to intergenic regions.

**Supplementary Figure 2:**
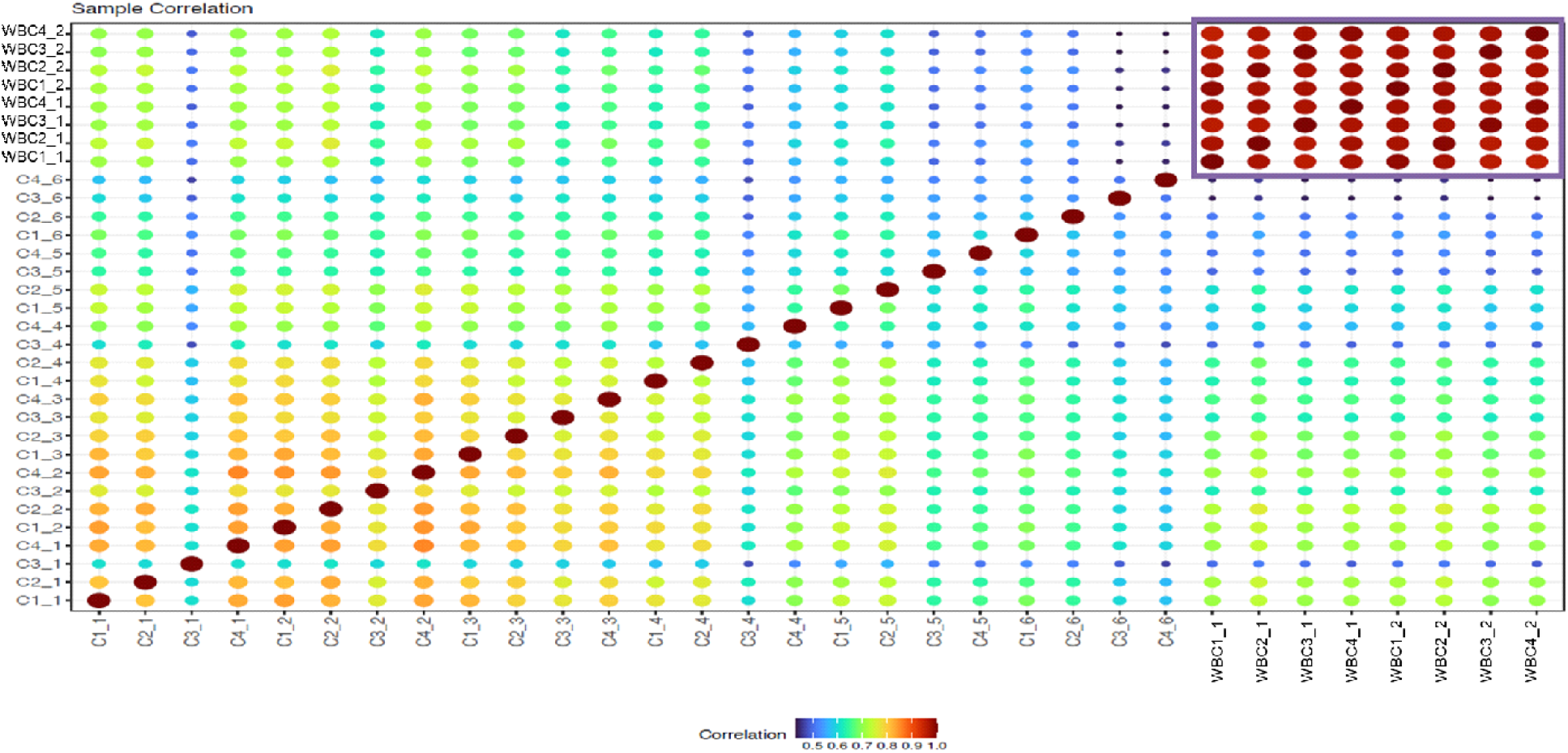
Between sample correlation of serially diluted cfRNA and cRNA isolated from the same whole blood sample. The dots represent correlation coefficients for abundance of transcripts between samples excluding those for hemoglobin, mitochondrial and rRNA genes. C1 to C4 represent the four cfRNA that were serially diluted; WBC_1 to WBC_4 represent the cRNA samples originating from the same individuals. The numbers _1 through _6 represent the six concentrations obtained from serial dilution of RNA samples with _1 being the undiluted sample.

## DISCUSSION

Next generation sequencing technologies for liquid biopsies have the potential to revolutionize cancer detection. While cfDNA has been widely studied, cfRNA is the next step in advancing cancer screening and treatment. cfRNA has the distinct potential to increase early detection of cancers yet to our knowledge few studies have attempted whole exome sequencing on cfRNA for cancer patients. Further, methods for cfDNA and cfRNA require large volumes of plasma, 5 mL or more, and often require specialized additives for collection tubes, which has the potential to limit availability of testing.(5, 9, 14).

In this study we chose to use samples obtained from standard 3-6 mL EDTA collection tubes routinely used in clinical care. The results of this study indicate volumes as low as 1.5 ml of EDTA plasma yielding less than 5ng of total cfRNA is sufficient for library preparation and sequencing. Our methodology produced acceptable sequence reads of 200 million to 350 million, with over 80% of reads mapped to the reference standard and >80% of those reads mapped to exonic regions, producing equivalent results to what is seen with methods requiring much high plasma starting volumes. However, the sequentially diluted cfRNA samples from healthy individuals demonstrated that the starting cfRNA concentration influenced the detection of differentially expressed genes. Whether these differences associated with smaller amounts of starting cfRNA are significant or relevant to detection of differentially expressed genes in lung cancer (or other cancers), will require a similar analysis on samples collected from those specific patient populations, ideally in studies specifically designed for acquiring larger amounts of samples. Another noteworthy finding is that the RNA in WBC and plasma isolated from the same blood sample yielded more than 8000 genes that were expressed differentially between the two tissues. These findings, despite the preliminary nature of our analyses, conform to previous reports that the cfRNA in plasma may provide a rich source of RNA that are not limited to those originating from blood cells.

Overall, there were no significant differences between cancer and non-cancer patients in terms of sample quality and mapping indicating our method for sample collection, storage, extraction, preparation, and sequencing is a viable and feasible option for cfRNA characterization. This proof-of-concept study provides a framework for screening cfRNA for identifying biomarkers for early detection of lung cancer (and other cancers), using minimal amounts of samples. However, this is only the first step in moving cfRNA liquid biopsies into the clinical arena. While this early study indicates use of small volumes has the potential to provide reproducible results, many parameters including precision, accuracy, limit of detection, sensitivity, and specificity, will need to be tested and standardized for adoption of this method into a CLIA-certified lab and FDA approval, which is needed for liquid biopsies to become standard of care(15, 16). Future work will require studies to identify cfRNA biomarkers, followed by rigorous testing to ensure these biomarkers can reliably be detected in patient populations and provide meaningful information that providers can act on to improve patient outcomes.

## Data Availability

All data produced in the present study are available upon reasonable request to the authors

## AKNOWLEDGEMENTS

Authors would like to thank Dan Gheba and Tara Kesteloot (from Illumina) for their expert advice with developing the assays. Jessica Cornell and John Sivinski for their assistance with sample preparations and RNA extractions; Dr. Lisa Sadzewicz and Sandra Ott for advice and assistance with two-step sequencing.

## REFERENCES

1. Bade BC, Cruz CSD. Lung cancer 2020: epidemiology, etiology, and prevention. Clin. Chest Med. 2020;41(1):1–24.

2. Rahib L, Wehner MR, Matrisian LM, Nead KT. Estimated Projection of US Cancer Incidence and Death to 2040. JAMA Netw. Open. 2021;4(4):e214708–e.

3. Latimer K, Mott T. Lung cancer: diagnosis, treatment principles, and screening. Am Fam Physician. 2015;91(4):250–6.

4. Arrieta O, Villarreal-Garza C, Martínez-Barrera L, Morales M, Dorantes-Gallareta Y, Peña-Curiel O, et al. Usefulness of serum carcinoembryonic antigen (CEA) in evaluating response to chemotherapy in patients with advanced non small-cell lung cancer: a prospective cohort study. BMC cancer. 2013;13(1):1–7.

5. Ilié M, Hofman P. Pros: Can tissue biopsy be replaced by liquid biopsy? Transl Lung Cancer Res. 2016;5(4):420.

6. Drag MH, Kilpeläinen TO. Cell-free DNA and RNA—measurement and applications in clinical diagnostics with focus on metabolic disorders. Physiol. Genomics. 2021;53(1):33–46.

7. Russano M, Napolitano A, Ribelli G, Iuliani M, Simonetti S, Citarella F, et al. Liquid biopsy and tumor heterogeneity in metastatic solid tumors: the potentiality of blood samples. J. Exp. Clin. Cancer Res. 2020;39:1–13.

8. Gerlinger M, Rowan AJ, Horswell S, Larkin J, Endesfelder D, Gronroos E, et al. Intratumor heterogeneity and branched evolution revealed by multiregion sequencing. N Engl j Med. 2012;366:883–92.

9. Larson MH, Pan W, Kim HJ, Mauntz RE, Stuart SM, Pimentel M, et al. A comprehensive characterization of the cell-free transcriptome reveals tissue-and subtype-specific biomarkers for cancer detection. Nat. Commun. 2021;12(1):1–11.

10. Chan KA, Jiang P, Zheng YW, Liao GJ, Sun H, Wong J, et al. Cancer genome scanning in plasma: detection of tumor-associated copy number aberrations, single-nucleotide variants, and tumoral heterogeneity by massively parallel sequencing. Clin. Chem. 2013;59(1):211–24.

11. Chen M, Zhao H. Next-generation sequencing in liquid biopsy: cancer screening and early detection. Hum. Genomics. 2019;13(1):1–10.

12. Kopreski MS, Benko FA, Kwak LW, Gocke CD. Detection of tumor messenger RNA in the serum of patients with malignant melanoma. Clin. Cancer Res. 1999;5(8):1961–5.

13. Leng Q, Lin Y, Jiang F, Lee CJ, Zhan M, Fang H, Wang Y, Jiang F. A plasma miRNA signature for lung cancer early detection. Oncotarget. 2017;8(67).

14. Sorber L, Zwaenepoel K, Jacobs J, De Winne K, Goethals S, Reclusa P, et al. Circulating cell-free DNA and RNA analysis as liquid biopsy: Optimal centrifugation protocol. Cancers. 2019;11(4):458.

15. Lu Y-T, Delijani K, Mecum A, Goldkorn A. Current status of liquid biopsies for the detection and management of prostate cancer. Cancer Manag. Res. 2019;11:5271.

16. Goodsaid FM. The labyrinth of product development and regulatory approvals in liquid biopsy diagnostics. Clin Transl Sci. 2019;12(5):431–9.

